# Dominant harmonic pattern as an ictal marker of the epileptogenic zone in focal neocortical epilepsy

**DOI:** 10.1101/2024.09.25.24314351

**Authors:** Lingqi Ye, Lingli Hu, Hongyi Ye, Yihe Chen, Junming Zhu, Zhe Zheng, Hongjie Jiang, Dongping Yang, Cong Chen, Shan Wang, Zhongjin Wang, Wenjie Ming, Yi Wang, Cenglin Xu, Jin Wang, Meiping Ding, Shuang Wang

## Abstract

**Objective:** The ictal *Harmonic* pattern (*H* pattern), produced by the non-linear characteristics of EEG waveforms, may hold significant potential for localizing the epileptogenic zone (EZ) in focal epilepsy. However, further validation is needed to establish the *H* pattern’s effectiveness as a biomarker for measuring the EZ.

**Methods:** We retrospectively enrolled 131 patients diagnosed with drug-resistant focal epilepsy, all of whom had complete stereo-electroencephalographic (SEEG) data. From this cohort, we selected 85 patients for outcome analysis. We analyzed the morphological and time-frequency (TF) features of the *H* pattern using TF plots. A third quartile (Q3) threshold was applied to classify channels expressing either dominant (*Channel_dH pattern_*) or non-dominant *H* patterns (*Channel_non-dH pattern_*). We then examined associations between the morphological features of the *H* pattern and patients’ clinical characteristics, as well as the correlations between the extent of channel removal and seizure outcomes.

**Results:** We found no significant correlations between the morphological features of the ictal *H* pattern and clinical factors, including lesional MRI findings, epileptic onset patterns, epilepsy type, pathology, or surgical outcomes. The non-localizable *H* pattern appeared exclusively in patients with non-focal onset patterns. Notably, the proportion of *Channel_dH pattern_* was higher in the seizure-onset zone (SOZ) compared to the early propagation zone. The seizure-free group demonstrated significantly higher removal proportions of *Channel_dH pattern_*, both within and outside the SOZ (*p* = 0.014; *p* = 0.036), with AUCs of 0.606 and 0.660, respectively, in a seizure freedom prediction model. Survival analysis confirmed that complete removal of these regions correlated with long-term seizure freedom (*p* = 0.008; *p* = 0.028). Further subgroup analysis showed a significant correlation in neocortical epilepsy (*p* = 0.0004; *p* = 0.011), but not in mesial temporal lobe epilepsy. Additionally, multivariate analysis identified the complete removal of *Channel_dH pattern_* as the only independent predictor for seizure freedom (*p* = 0.022; OR 6.035, 95% CI 1.291-28.211).

**Conclusions:** Our study supports the notion that the dominance of the ictal *H* pattern, regardless of its morphology, serves as a novel biomarker for the EZ in focal epilepsy. The non-linearity in EEG waveforms provides new insights into understanding ictal spreading propagation and offers potential improvements for surgical planning in neocortical epilepsy.

## Introduction

Resective surgery currently remains the most effective treatment for drug-resistant epilepsy (DRE). However, only 50% of patients achieve prolonged seizure freedom following surgery. Despite advancements in intracranial electroencephalography (iEEG) technology, it has yet to surpass a 70% ceiling in achieving favorable surgical outcomes ^1–3^. The success of the surgery largely depends on the precise localization and complete removal of the epileptogenic zone (EZ), which is defined as the brain area essential for generating seizures ^4^. Postsurgical seizure freedom indicates that the EZ was likely successfully removed. Although the seizure-onset zone (SOZ) does not necessarily equate to the EZ ^5, 6^, experienced clinicians’ visual identification of the SOZ is a crucial step in the process of localizing the EZ.

Many studies have aimed to identify ictal EEG biomarkers capable of quantitatively delineating the EZ. Established semi-quantitative algorithms, such as the epileptogenic index (EI) ^7^ and epileptogenic map ^8^, have been shown to be effective in cases where ictal onset patterns include low-voltage fast activity. Additionally, the removal of areas exhibiting ictal high-frequency oscillations (HFOs) has been reported as a prognostic indicator of favorable seizure outcomes ^9–11^. The concept of the EEG *fingerprint*, proposed as an ictal biomarker of the EZ, suggests a possible synchronous activation of GABAergic neurons at seizure onset ^12^. Similarly, *brain chirp* ^13^ highlights the importance of fast EEG activity in defining the EZ.

In our previous study, we identified the *Harmonic* pattern (*H* pattern), a spectral feature of specific ictal EEG components that contains valuable information for seizure propagation and EZ localization. The *H* pattern is characterized by an equidistant spectral band distribution of varying frequencies on time-frequency (TF) plots. Generated by the specific non-sinusoidal waveforms, the ictal *H* pattern may represent periodically synchronized neural firings, rather than a methodological artifact. Furthermore, the *H* pattern is frequently observed in seizures with various onset patterns and across different seizure stages ^14^. The dominance of the ictal *H* pattern in different brain regions reflects higher non-linearity and has proven useful in localizing the EZ.

Despite its potential, the full spectrum of the ictal *H* pattern remains unclear. The *H* pattern shows significant variability in morphology across patients; however, within an individual patient, its morphology and spatial localization tend to remain consistent across seizures, emphasizing the personalized nature of ictal neural dynamics. In this expanded cohort, we first investigated clinical factors related to the morphological properties of the *H* pattern, specifically focusing on its clarity and shape. High clarity indicates a narrower frequency band with reduced background noise, while the shape of the spectral bands reflects changes in the frequency of ictal neural oscillations at a macroscopic level. This study aims to further support the hypothesis that the dominance of the ictal *H* pattern could serve as a novel biomarker for the EZ. Additionally, we analyzed the potential impact of the *H* pattern’s morphological features.

## Methods

### Selection of patient cohort

We retrospectively reviewed 133 patients with drug-resistant epilepsy who had completed stereo-electroencephalography (SEEG) at the Second Affiliated Hospital Zhejiang University School of Medicine (SAHZU) between April 2013 and October 2022. We excluded two patients diagnosed with hypothalamic hamartoma from the study. Therefore, we performed *H* pattern analysis on the remaining 131 cases. From this cohort, we selected 85 patients for outcome analysis based on the following inclusion criteria (**Figure S1**): (1) a focal-onset pattern on SEEG with a clearly defined SOZ localized to a discrete cortical region; (2) undergoing either resective surgery or thermocoagulation treatment; (3) recording at least two habitual clinical seizures during SEEG monitoring; (4) identifying a discernible and localizable ictal *H* pattern (as described in the *Identification of ictal H pattern* section); (5) a follow-up period of at least 12 months after surgery. The Medical Ethics Committee of SAHZU approved this study (Study No. 2023-1260, 2021-0585). We obtained written informed consent from each participant.

### SEEG evaluation

All patients were recommended for SEEG evaluation during a multidisciplinary patient management conference. The non-invasive presurgical evaluation included a detailed medical history, neuropsychological assessment, scalp video-EEG monitoring, 3.0-Tesla cerebral MRI, and interictal ^18^F-fluorodeoxyglucose positron emission tomography (^18^F-FDG-PET). The procedure for SEEG electrode implantation has been previously described ^15^. Based on the working hypothesis, multiple-contact intracerebral electrodes (8-16 contacts, each 2 mm in length, 0.8 mm in diameter, and spaced 1.5 mm apart) were implanted using a robotic arm-assisted system. SEEG signals were recorded at sampling rates of either 1,000 or 2,000 Hz using EEG systems with 256 channels (Nihon Kohden) or 128 channels (Xltek, Natus). The anatomical locations of each contact along the electrode trajectory in all subjects were confirmed by co-registering post-implantation CT scans with pre-implantation MRI using *Sinovation 2.0* ^16^. A three-dimensional schematic diagram of the reconstruction is provided in the flowchart (**Figure 1C**).

**Figure 1.**
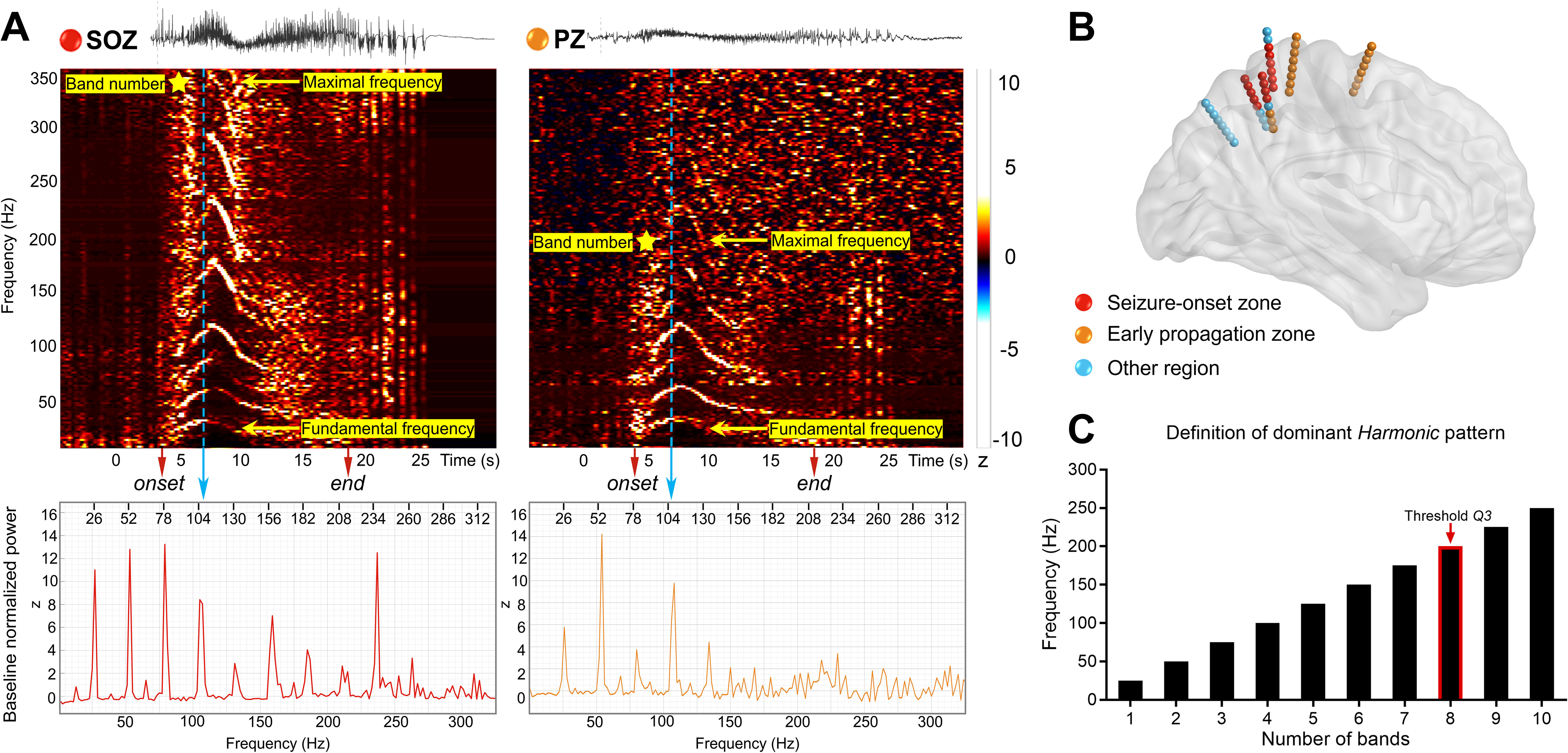
Examples and characteristics of the *H* pattern (data from patient #057). (A) Ictal SEEG recordings from both SOZ and PZ of a patient with right parietal focal cortical dysplasia showing bursts of high-amplitude poly-spikes followed by low-voltage fast activity at seizure onset. Below the SEEG recordings are corresponding time-frequency plots of the two channels. An ictal *H* pattern could be identified in both plots: two or more unique high-intensity spectral bands with varying frequency starting at 4 s (red arrow) after 0 s (seizure onset), and ending at 19 s (red arrow). The frequencies of the bottom and top spectral bands are defined as the *fundamental frequency* and *maximal frequency*, respectively (yellow arrow). Importantly, the ictal *H* pattern exhibits a distinctive feature of equidistant distribution on the TF plots. For instance, at 6.3 s (long blue dotted arrows), the frequency interval between adjacent spectral bands is nearly uniform, at approximately 26-28 Hz, as observed in the power spectral density diagrams. The ictal *H* pattern shown in the two channels are classified as a ‘dominant *H* pattern (d*H* pattern)’ and a ‘non-dominant *H* pattern (non-d*H* pattern)’, respectively. (B) 3D representation of an example SEEG exploration. Electrode contacts are categorized into three regions: the SOZ (red dots), the PZ (orange dots), and the other regions (blue dots), according to a patient’s ictal SEEG recordings. (C) Total number of visually determined spectral bands on the TF plot of each channel each time a seizure is counted. Channels are then ranked based on the number of bands, and if a channel’s count exceeds the threshold of *Q3*, it is marked as expressing the ictal dominant *H* pattern. SEEG, stereo-electroencephalography; SOZ, seizure-onset zone; PZ, early propagation zone; *H* pattern, *Harmonic* pattern; *Q3*, the 75th percentile of data distribution.

### Surgical outcome

Following SEEG evaluation, patients underwent either resective surgery or laser ablation. The planning of these surgical interventions was conducted independently of this study. Each contact was classified as either *removed* or *non-removed* based on postsurgical MRI evaluations. Surgical outcomes were assessed at the most recent follow-up and patients were categorized as either seizure-free (SF, Engel class Ia) or not seizure-free (NSF, >Engel class Ia) ^17^.

### Visual EEG analysis

Two experienced and board-certified neurophysiologists reviewed the SEEG data to identify the SOZ and the rapid propagation zone (PZ) using our previously established methods ^15, 18^. The SOZ is defined as the brain area that shows the earliest changes on SEEG. The PZ is characterized by the rapid spread of ictal activity within 3-5 seconds after seizure onset. Areas outside these defined zones were classified as ‘other regions.’ Both the PZ and these ‘other regions’ were collectively categorized as non-SOZ.

### Identification of ictal H pattern

We selected only contacts within cortical gray matter for quantitative analysis. We analyzed all EEG data using a bipolar montage. For each identified seizure, we extracted a 110-second segment of SEEG data: 10 seconds before and 100 seconds after the EEG onset. Additionally, we collected a segment of preictal EEG data ranging from −230 to −120 seconds before EEG onset as baseline data for normalization. To enhance the reliability of our results, we applied two analytical methods: the Morlet wavelet transform^19^ and the multitaper method ^20, 21^ in the TF analysis.

We identified the ictal *H* pattern from TF maps based on the following criteria: the presence of two or more distinct high-intensity bands with varying frequencies occurring at any point during a seizure, with equal frequency intervals between adjacent bands (**Figure 1A**). For each patient, we analyzed only the initially occurring *H* pattern. We estimated various features of the ictal *H* pattern, including its shape, clarity, onset and end times, fundamental and maximal frequencies, frequency intervals, and the number of bands. If the *H* pattern was consistently stereotyped across different seizures for a single patient, we included only one data set in the analysis. To determine the dominance of the *H* pattern, we established a threshold known as *Q3*, which represents the upper quartile (75th percentile) of the data distribution for the maximal count of frequency bands per patient. We calculated *Q3* using the following formula:

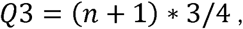

where *n* denotes the highest count of frequency bands observed across all channels. If the number of bands in a channel showing the ictal *H* pattern (*Channel_H pattern_*) exceeds the *Q3* threshold, we marked it as exhibiting a dominant *H* pattern (*Channel_dH pattern_*). Conversely, a channel not meeting this threshold was classified as expressing a non-dominant *H* pattern (*Channel_non-dH pattern_*) (**Figure 1D**). A localizable *H* pattern should be consistent across seizures and present in specific regions. If no gradient in band numbers was observed across all channels, the *H* pattern was classified as non-localizable.

### Morphology of Ictal Harmonic Pattern

We classified the clarity of the *H* pattern into two levels. Grade A indicated a highly clear frequency band with distinct borders against the background activity. Grade B referred to a less clear frequency band with fuzzier borders. We identified three distinct shapes of the *H* pattern: Shape 1 resembled a parabola, where the fundamental frequency initially increases and then decreases; Shape 2 appeared as a declining line, representing a decrease in the frequency harmonics of the spectral band; and Shape 3 took the form of an ascending line, indicating an increase in the frequency harmonics of the spectral band. This resulted in a total of six categories: A1, A2, A3, B1, B2, and B3 (**Figure S2**). Initially, two investigators (Ye L.Q. and Hu L.L.) performed these classifications. In cases of disagreement, a third epileptologist (Wang S.) made the final decision.

### Prediction of Surgical Outcome

For each patient, we categorized channels as either SOZ or non-SOZ based on visual analysis, and as *Channel_dH pattern_* or *Channel_non-dH pattern_* based on TF analysis. We determined the ratio of removed channels using the following formula:

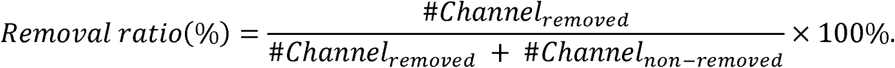

To estimate the correlation between the removal of *Channel_dH pattern_* and seizure freedom, we used the *Chi-Squared* test and receiver operating characteristic (ROC) curve analysis, from which we derived the area under the curve (AUC). We then generated Kaplan-Meier (KM) curves to depict the cumulative probability of achieving seizure freedom. We compared the retention of seizure freedom between groups using the log-rank test, including patients with complete (100%) removal and those with incomplete or no removal (0-100%).

### Statistical Analysis

We used MATLAB 2021a (The MathWorks, Inc.) and SPSS 26.0 for statistical analysis. We evaluated the normality of the distribution of all continuous variables using the *Kolmogorov-Smirnov* test. Normally distributed variables were presented as mean ± standard deviation (SD), while non-normally distributed variables were reported as median (interquartile range, IQR). We expressed categorical variables as frequencies. We applied an *unpaired t*-test or a nonparametric *Mann-Whitney U* test to analyze continuous variables, and we assessed categorical variables using the *Chi-Squared* test. We also conducted a binary regression analysis to identify predictive factors affecting surgical outcomes. For multiple comparisons, we employed the Bonferroni correction method. *p* < 0.05 represents statistically significant.

## Results

### Patient Demographics

In the entire cohort, 75.6% (99 patients) exhibited a focal-onset pattern, while the remaining 24.4% (32 patients) had a non-focal onset pattern (**Table S1**). Among those with a focal-onset pattern, 92.9% (92 patients) displayed an ictal *H* pattern, all of which were localizable. In contrast, 93.8% (30 patients) of those with a non-focal onset pattern exhibited at least one ictal *H* pattern. Of these, 80.0% (24 patients) were localizable, and 20.0% (6 patients) were non-localizable. Most patients presented with only one type of *H* pattern. However, two different types of *H* pattern were observed exclusively in approximately 20% of patients with a non-focal onset pattern (**Table S1**).

Of the 85 patients included in the outcome analysis, 38.9% (33 patients) had participated in our previous *H* pattern study (**Table 1**) ^14^. In the current cohort, 61.2% (52 patients) achieved seizure freedom after an average follow-up period of 47.8 months. We found no significant differences between the SF and NSF groups regarding age at disease onset, age at SEEG recording, or the lateralization/localization of seizures. The incidence of detectable abnormalities on MRI was similar between the two groups (75.0% in the SF group and 72.7% in the NSF group, *p* = 0.816). However, the NSF group had a significantly higher number of implanted electrodes (*p* = 0.025). Focal cortical dysplasia (FCD) LJ was the most common pathological finding, occurring in 34.1% of patients. Among those with malformations of cortical development (MCD) other than FCD LJ (19.5% of patients), 11 had FCD I, 3 had tuberous sclerosis complex, 1 had FCD LJc, and 1 had a mild malformation of cortical development with oligodendroglial hyperplasia and epilepsy. Patients with FCD LJa were included in the hippocampal sclerosis (HS) group. We observed a significant difference in the pathological findings between the SF and NSF groups (*p* = 0.014), particularly between FCD LJ and gliosis/non-specific findings.

**Table 1.** Patients’ demographics and surgical outcome.

|  | Total cohort | Seizure-free<br>(Engel class Ia) | Non-seizure free<br>(> Engel class Ia) | *p* value |
| --- | --- | --- | --- | --- |
| Number of patients, n | 85 | 52 | 33 | - |
| Gender, M, n (%) | 47 (55.3%) | 29 (55.8%) | 18 (54.5%) | 0.912<sup>a</sup> |
| Age at onset, y | 10.0 (6.0-15.4) | 8.5 (5.8-18.8) | 12.0 (6.5-15.4) | 0.338<sup>c</sup> |
| Age at SEEG recording, y | 23.3 ± 10.4 | 23.0 ± 11.1 | 23.7 ± 9.4 | 0.753<sup>b</sup> |
| EEG pattern at H pattern, n (%) |  |  |  |  |
| Irregular spiking (IS) | 29 (34.1%) | 17 (32.7%) | 12 (36.4%) | 0.728<sup>a</sup> |
| Fast activity (FA) | 56 (65.9%) | 35 (67.3%) | 21 (63.6%) |  |
| Lateralization, n (%) |  |  |  |  |
| Left | 36 (42.4%) | 21 (40.4%) | 18 (54.5%) | 0.645<sup>a</sup> |
| Right | 49 (57.6%) | 31 (59.6%) | 15 (45.5%) |  |
| Localization, n (%) |  |  |  |  |
| Mesial temporal lobe | 28 (32.9%) | 15 (28.8%) | 13 (39.4%) | 0.313<sup>a</sup> |
| Neocortex | 57 (67.1%) | 37 (71.2%) | 20 (60.6%) |  |
| Lesional MRI, n (%) | 63 (74.1%) | 39 (75.0%) | 24 (72.7%) | 0.816<sup>a</sup> |
| Number of Electrodes, n | 10.0 (8.0-11.5) | 9.0 (7.0-11.0) | 10.0 (9.0-12.5) | **0.025<sup>c</sup>** |
| Number of Contacts, n | 112.0 (88.5-127.0) | 112.5 (81.3-127.8) | 112.0 (96.0-126.5) | 0.433<sup>c</sup> |
| Bilateral implantation, n (%) | 27 (31.8%) | 16 (30.8%) | 11 (33.3%) | 0.874<sup>a</sup> |
| Pathology, n (%) | 82 | 52 | 30 |  |
| Hippocampal sclerosis | 13 (15.9%) | 8 (15.4%) | 5 (16.7%) | **0.014<sup>d</sup>** |
| FCD type-II | 28 (34.1%) | 22 (42.3%)<sup>e</sup> | 6 (20.0%)<sup>e</sup> |  |
| Other MCD types | 16 (19.5%) | 11 (21.2%) | 5 (16.7%) |  |
| Tumor | 3 (3.7%) | 3 (5.8%) | 0 |  |
| Gliosis/other non-specific | 22 (26.8%) | 8 (15.4%)<sup>f</sup> | 14 (46.7%)<sup>f</sup> |  |

Normal variables were presented as mean ± standard deviation (SD). Non-normal variables were reported as median (interquartile range, IQR). a. *Pearson's chi-square* test. b. *Unpaired t-test*. c. *Mann-Whitney U* test. d. *Fisher exact* test. e & f. *Bonferroni* correction: In the SF and NSF groups, there was a discrepancy in pathological finding ( $p = 0.014$ ). A significant difference was found between FCD type-II and gliosis/non-specific findings.
SEEG, stereo-electroencephalography, H pattern, Harmonic pattern, MRI, magnetic resonance imaging, FCD, focal cortical dysplasia, MCD, malformation of cortical development
Notion: The category *Other MCD types* consists of FCD type-I, tuberous sclerosis complex, heterotopia, and mild malformation of cortical development with oligodendroglial hyperplasia in epilepsy.

### Morphology, Distribution and Parameters of Ictal Harmonic Pattern

We summarized the morphological characteristics of the ictal *H* pattern in patients who exhibited only one type of *H* pattern in **Table S2**. The most frequently observed shapes were the declining line (57.3%) and the parabola (40.2%) (**Figure S2**). Our analysis revealed no correlation between the clarity or shape of the *H* pattern and clinical factors, including the presence of lesional MRI findings, focal-onset pattern, neocortical epilepsy, pathology, or surgical outcome.

Out of 85 patients with focal-onset patterns, we identified both the SOZ and the PZ in 65 patients. Within these zones, the proportion of channels exhibiting the ictal *H* pattern was significantly higher in the SOZ (100.0%, IQR 57.3-100%) compared to the PZ (26.3%, IQR 0-50.0%; *p* < 0.0001) (**Figure 2A**). Similarly, the ictal d*H* pattern appeared more frequently in the SOZ (38.5%, IQR 16.0-80.0%) than in the PZ (0%, IQR 0-5.8%; *p* < 0.0001) (**Figure 2A**). The ictal *H* pattern was less common in other regions, and we did not observe a significant difference in the distribution of the non-d*H* pattern between the SOZ and PZ. Among the 65 patients, 37 exhibited the ictal *H* pattern simultaneously in both the SOZ and PZ. The number of frequency bands of the ictal *H* pattern was notably higher in the SOZ (3.1, IQR 2.2-6.2) compared to the PZ (2.5, IQR 1.9-4.2; *p* < 0.0001), as was the maximal frequency (SOZ, 118.8, IQR 92.5-333.0 Hz; PZ, 90.0, IQR 66.3-112.6 Hz; *p* < 0.0001). The onset time of the ictal *H* pattern did not differ between the SOZ and PZ, indicating almost simultaneous initiation across these regions. Furthermore, we did not observe differences in the termination, fundamental frequency, or frequency interval of the ictal *H* pattern between the SOZ and PZ (**Figure 2B-D**). These parameters of the ictal *H* pattern also did not differ between the SF and NSF groups (**Figure 3B-E**). In the remaining 20 patients without a PZ, the d*H* pattern was predominantly expressed in 88.9% of channels within the SOZ (IQR, 54.6-88.9%) and was rarely observed in other regions.

**Figure 2.**
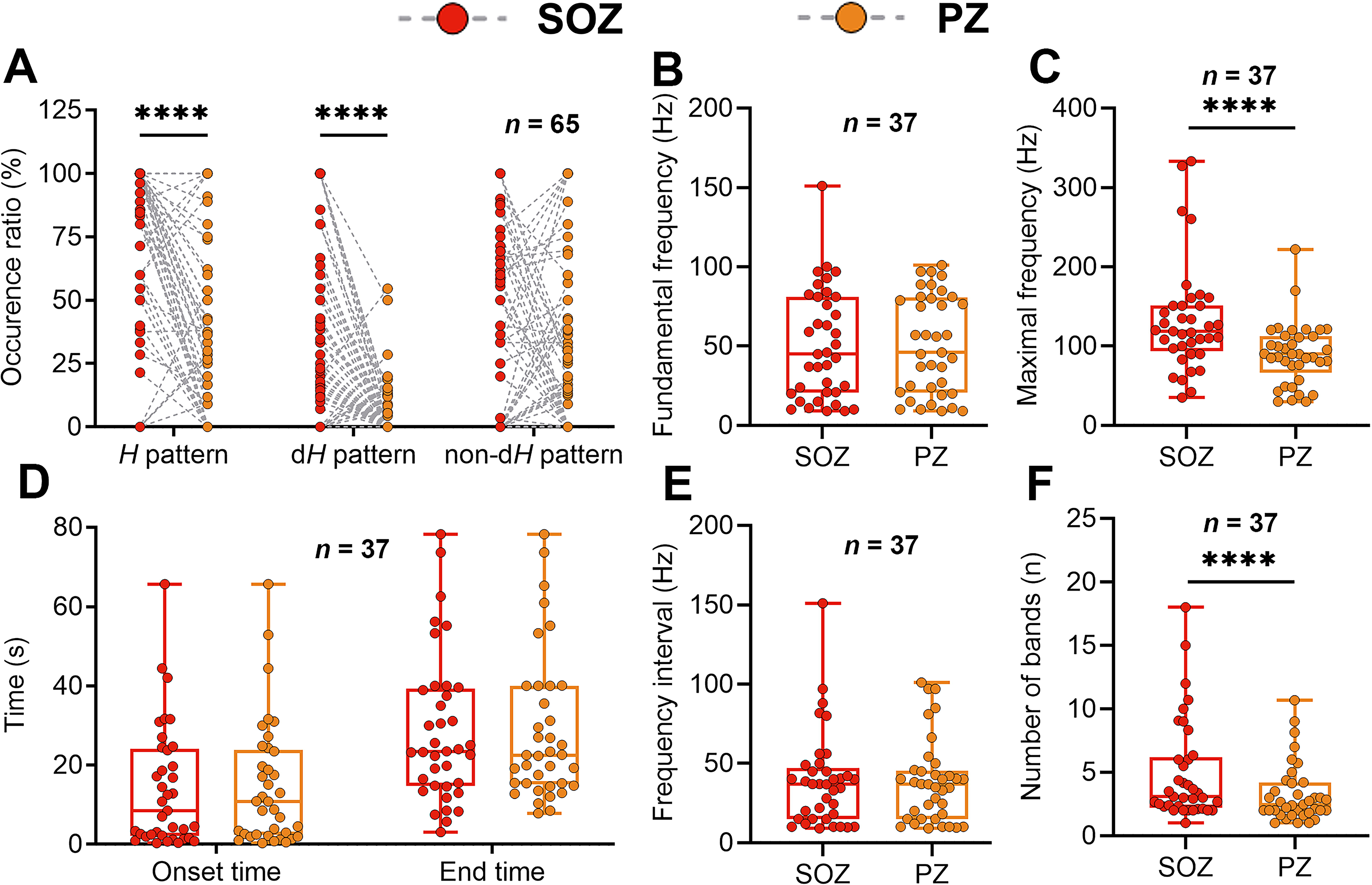
Distribution and time-frequency features of ictal *H* patterns in SOZ and PZ. (A) Proportions of channels expressing the ictal *H* pattern, the ictal dominant *H* pattern, and the ictal non-dominant *H* pattern between SOZ (red dots) and PZ (orange dots) in 65 patients who had both SOZ and PZ. (B-F) Onset and end time, fundamental frequency, maximal frequency, frequency interval, and number of spectral bands of the ictal *H* pattern between SOZ and PZ in 37 patients that simultaneously expressed the ictal *H* pattern in both SOZ and PZ. ****: *p* < 0.0001, nonparametric Mann-Whitney U test; *H* pattern, *Harmonic* pattern; SOZ, seizure-onset zone; PZ, early propagation zone.

**Figure 3.**
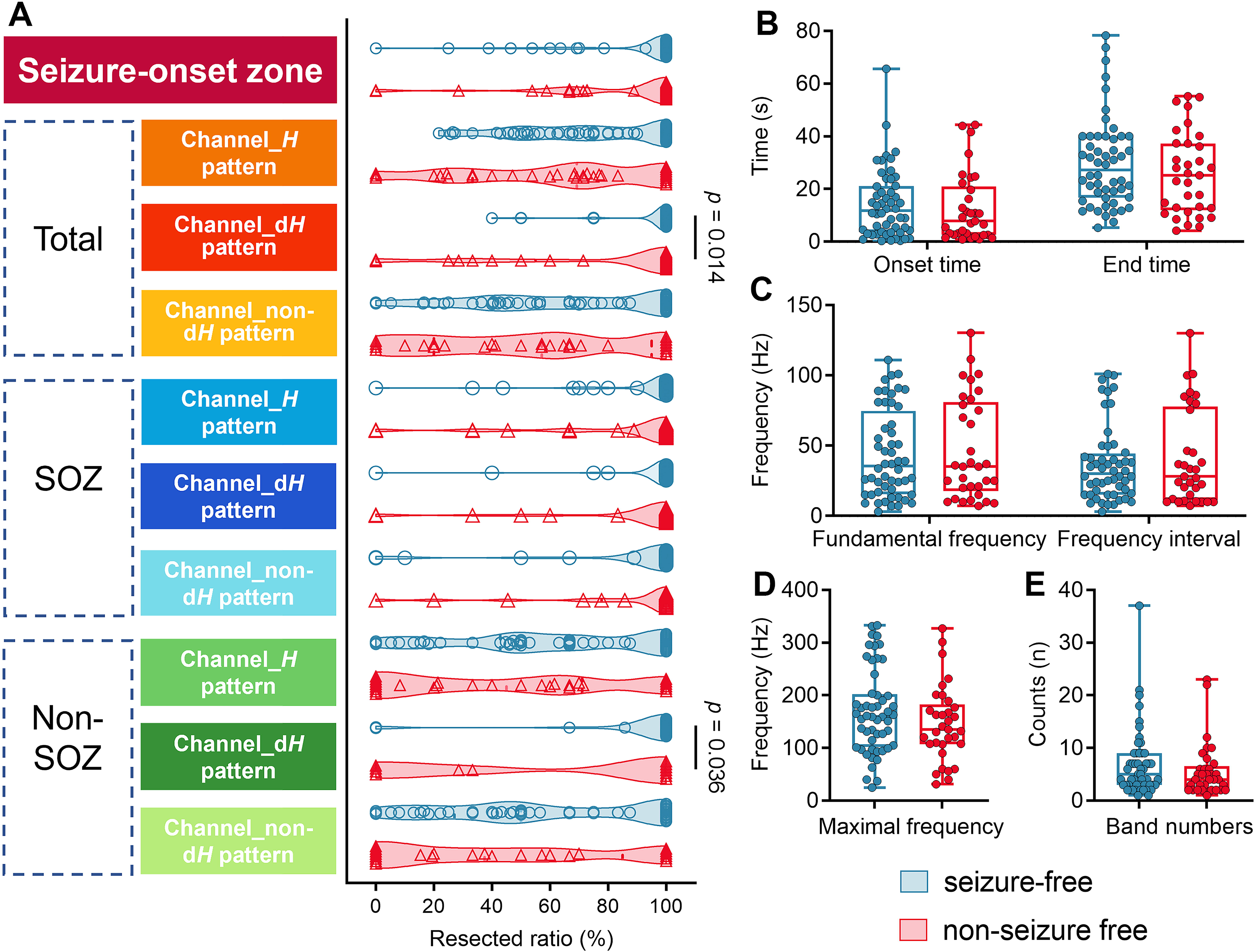
Associations between proportions of removal, time-frequency features of the ictal *H* patterns, and seizure outcomes. (A) Violin diagrams comparing removed ratios in channels of SOZ, channels expressing the ictal *H* pattern, channels expressing the ictal dominant *H* pattern, and channels expressing the ictal non-dominant *H* pattern between seizure-free (blue hollow circles) and not seizure-free (red hollow triangles) patients. (B-E) Onset time, end time, fundamental frequency, maximal frequency, frequency interval, and the number of spectral bands of the ictal *H* pattern between seizure-free (blue dots) and not seizure-free (red dots) patients. *H pattern*, *Harmonic* pattern; SOZ, seizure-onset zone.

### Ictal Dominant Harmonic Pattern

**Figure 3A** shows that a higher removal ratio of the *Channel_dH pattern_* significantly correlates with achieving seizure freedom (*p* = 0.014). The SF group demonstrated a higher removal ratio of the *Channel_dH pattern_* within non-SOZ (100.0, IQR 100-100%) compared to the NSF group (100.0, IQR 0-100%; *p* = 0.036).

### Predictive Value of Removing Ictal Dominant Harmonic Pattern

Regression analysis identified several independent factors significantly associated with achieving seizure freedom. These factors include the complete removal of the *Channel_dH pattern_* (*p* = 0.022; OR 6.035, 95% CI 1.291-28.211), FCD LJ or other MCD types (as opposed to gliosis or other non-specific pathologies), and a smaller number of implanted electrodes (**Figure 4**). Additionally, the association between the complete removal of the *Channel_dH pattern_* and seizure freedom was supported by an analysis showing statistical significance (*p* = 0.031, AUC = 0.606) (**Figure 5**). The highest AUC value of 0.660 was recorded when considering the complete removal of the *Channel_dH pattern_* within non-SOZ, highlighting the importance of the ictal d*H* pattern for localizing the EZ (**Figure S3)**.

**Figure 4.**
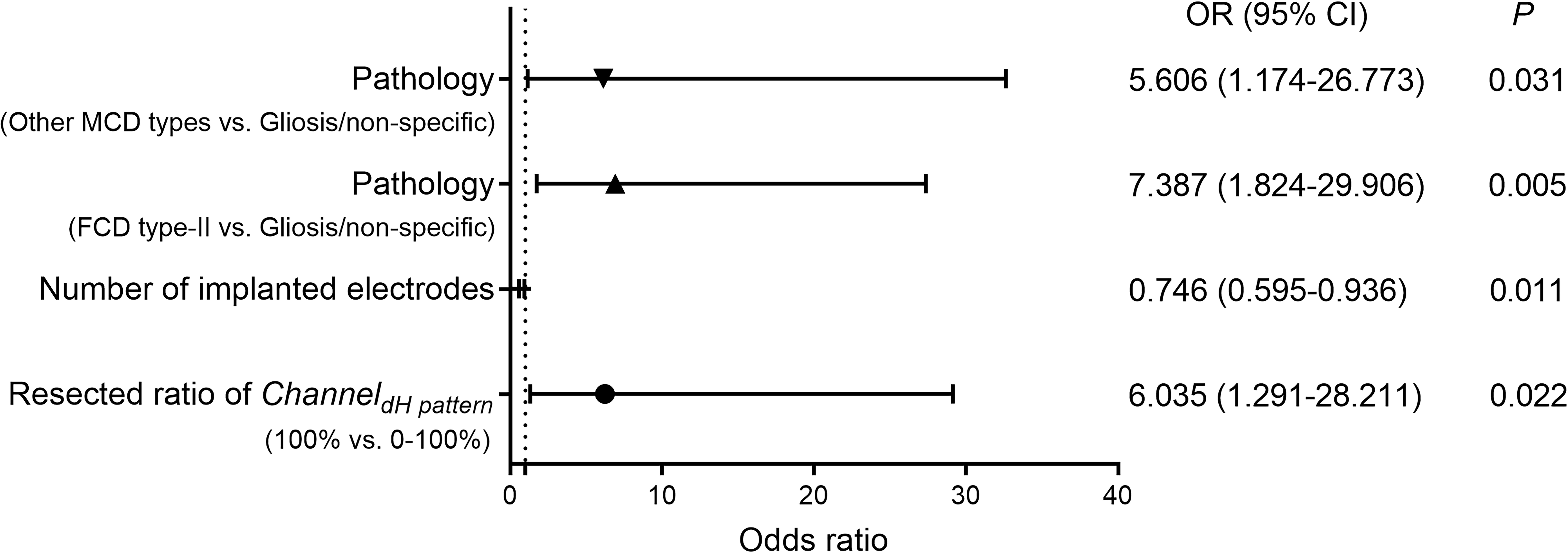
Binary regression for classification of seizure outcomes. Seven variables were recruited into the equation: EEG pattern harboring the *H* pattern, lesional MRI findings, seizure localization (mesial temporal lobe or neocortex), pathological findings, removed ratios of channels of the seizure-onset zone, channels expressing the ictal *H* pattern, and channels expressing the ictal dominant *H* pattern. The forest plot shows complete removal of channels expressing the ictal dominant *H* pattern, FCD type-II, or other MCD types (compared to gliosis/other non-specific pathology). Less implanted electrodes were correlated with seizure freedom. EEG, electroencephalography; *H* pattern, *Harmonic* pattern; MRI, magnetic resonance imaging; FCD, focal cortical dysplasia; MCD, malformation of cortical development; OR, odds ratio; CI, confidence interval.

**Figure 5.**
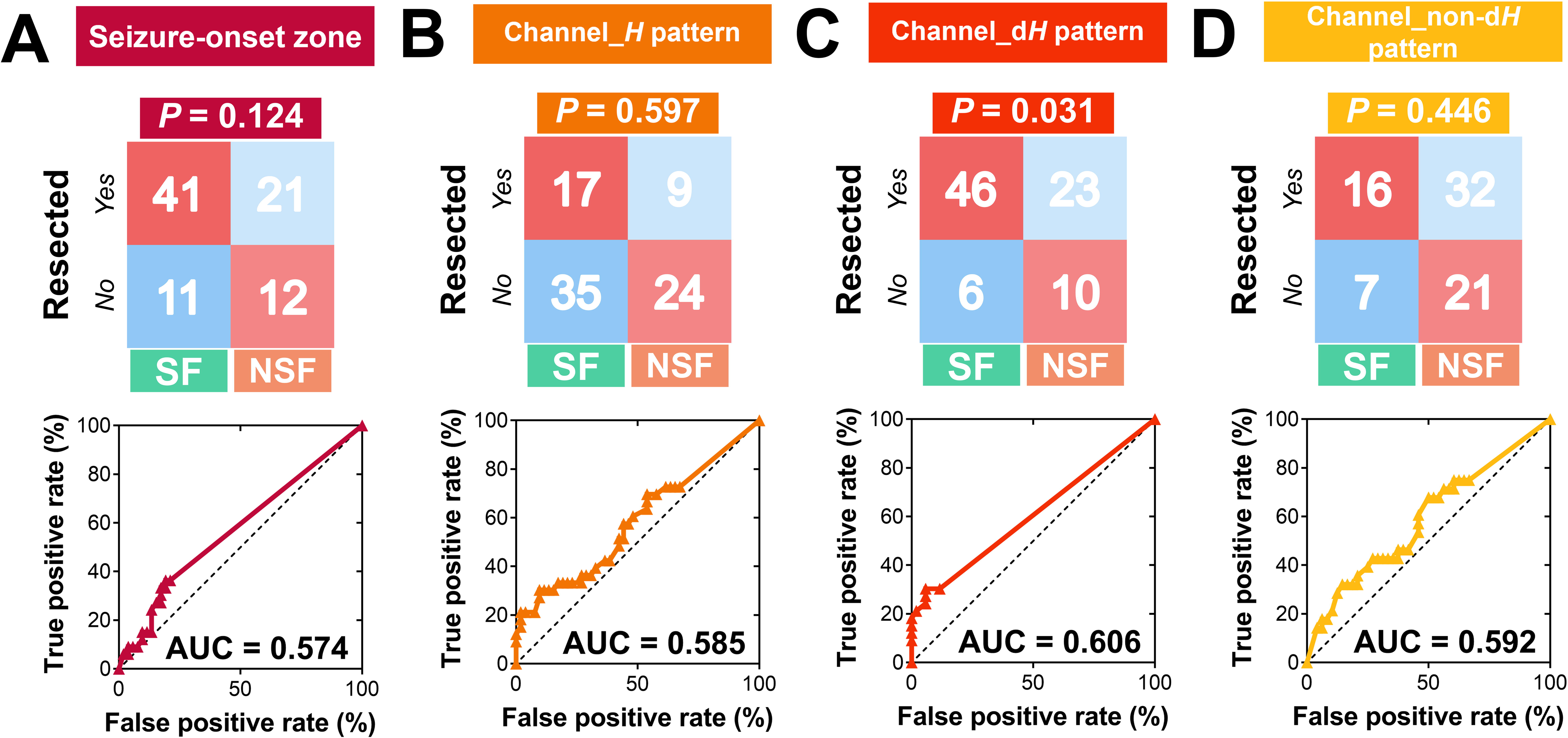
Predictive values for seizure outcomes. (A-D) Four-fold tables and receiver operator characteristic curve (ROC) for the removed ratios of channels within the seizure-onset zone (dark red), channels expressing the ictal *H* pattern (orange), channels expressing the ictal dominant *H* patten (scarlet), and channels expressing the ictal non-dominant *H* pattern (yellow) as predictors of post-surgical outcomes. In the four-fold tables, the numbers of seizure-free patients and not seizure-free patients with all the corresponding channels removed (’*Yes*’, upper left of the table), or incompletely/none removed (’*No*’, bottom left of the table) are listed. *p*-values for each *Chi-Squared* test are provided above each table. In the ROC curves, each colored dot represents a combined value of the false positive rate (X-axis) and the true positive rate (Y-axis). The areas under curves (AUCs) are respectively listed on the bottom right corner.

### Survival Analysis

**Figure 6** and **Figure S4** illustrate the 10-year prognosis of seizure freedom for patients who underwent *complete removal* (100%) compared to those with *incomplete* or *no removal* (0∼100%). Among the 85 patients in the cohort, 58.2% achieved seizure freedom at the 10-year mark. Notably, patients who underwent complete removal of the *Channel_dH pattern_* had a significantly higher probability of attaining long-term seizure freedom (*p* = 0.008). Specifically, among the 37 patients with complete removal of the *Channel_dH pattern_* within non-SOZ, there was also an increased likelihood of achieving seizure freedom (*p* = 0.028). In cases of neocortical epilepsy, the complete removal of the *Channel_dH pattern_* was strongly associated with long-term seizure freedom (*p*=0.0004), regardless of whether the removal occurred within the SOZ (*p* = 0.013) or non-SOZ (*p* = 0.011) (**Figure S5**). In contrast, for mesial temporal lobe epilepsy (mTLE), complete removal of the SOZ, rather than the *Channel_dH pattern_*,was significantly associated with better long-term seizure freedom outcomes (*p* = 0.019).

**Figure 6.**
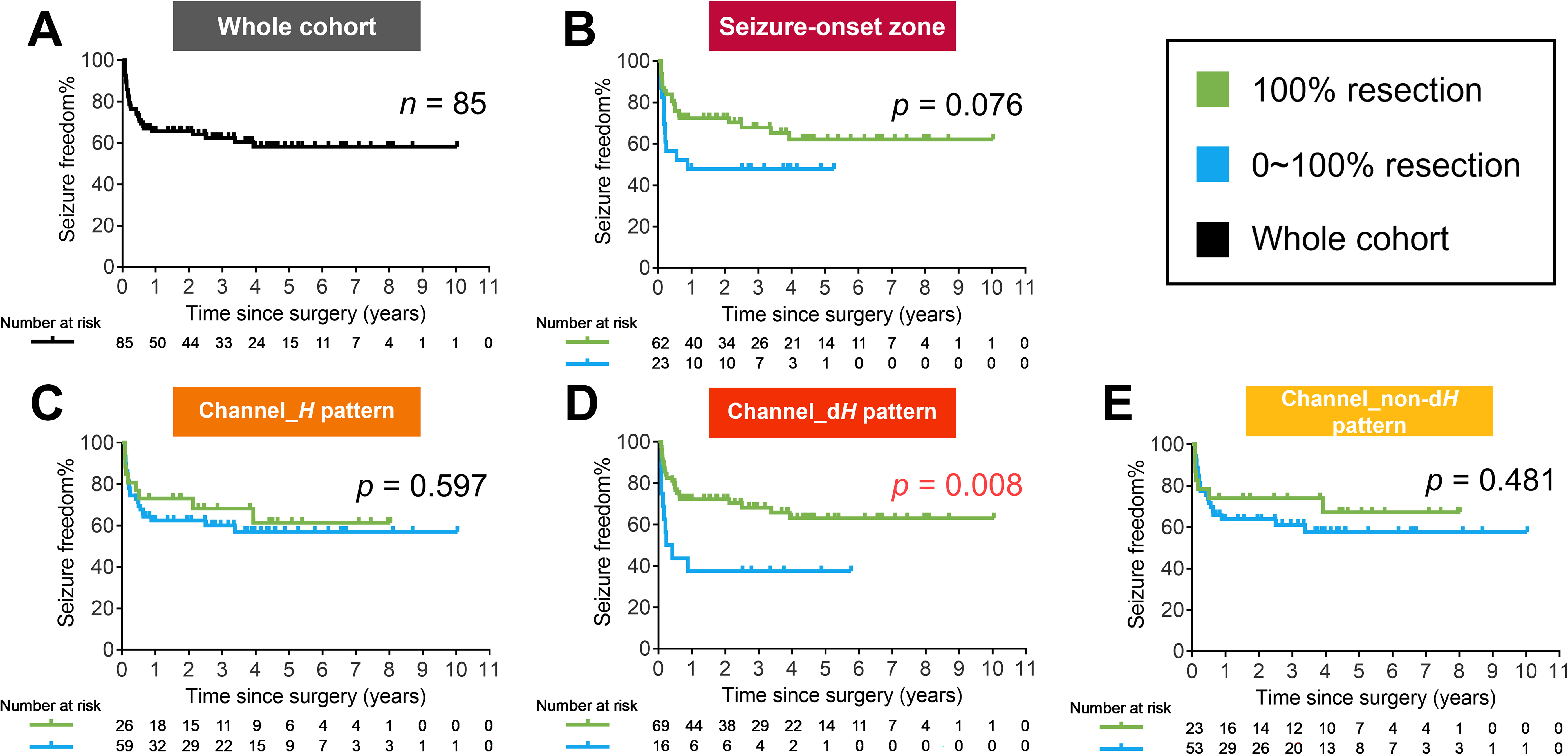
Survival analysis. Kaplan-Meier (KM) plots of cumulative probability of continuous seizure freedom separated by complete (100%) removal (green continuous line) and incomplete/none (0∼100%) removal (blue continuous line) of the whole cohort, channels of seizure-onset zone, channels expressing the ictal *H* pattern, channels expressing the ictal dominant *H* patten, and channels expressing the ictal non-dominant *H* pattern. The ‘number at risk (number censored)’ of each year is annotated below the KM plot for every category.

## Discussion

The ictal *H* pattern, which reflects the non-linear features of ictal SEEG signals on TF maps, may serve as an innovative ictal biomarker and offer a new approach for delineating the boundary of the EZ. Our key findings reveal: (1) In patients with focal onset patterns, the morphology of the ictal *H* pattern is consistent within individuals but heterogeneous across the cohort; (2) the morphological features of the ictal *H* pattern do not correlate with clinical variables; (3) the dominance of the ictal *H* pattern indicates high epileptogenicity, and removing regions that exhibit this pattern is crucial for achieving seizure freedom in focal epilepsy, particularly in neocortical epilepsy.

Previous studies have introduced various quantitative biomarkers for measuring spectral components of SEEG signals, including EI ^7, 22^, HFOs ^23^, and ictal non-linear metrics such as the *h*^2^ coefficient ^24, 25^, coherence analysis ^26, 27^, entropy ^28, 29^, and infraslow activity ^30–32^. These computational EEG markers aim to enhance the precision of EZ delineation. Our findings suggest that the ictal d*H* pattern could be a valuable biomarker for localizing the core epileptogenic cortex across different brain regions. Visual inspection showed that the distribution of the ictal *H* pattern aligned with the ictal onset zone, regardless of whether it was focal-, multifocal-, or diffuse-onset. In individuals with focal-onset patterns, the ictal *H* pattern typically exhibited a consistent spatial distribution across various seizures. In contrast, individuals with non-focal onset patterns displayed variable types of the ictal *H* pattern, indicating an unstable seizure propagation pattern. The near-simultaneous onset of the ictal *H* pattern across different regions, along with a uniform fundamental frequency, suggests clear inter-regional synchronization during this period. Notably, our findings indicate that regions showing the d*H* pattern can extend into the PZ beyond the boundary of the SOZ. As our previous research indicated, the *H* pattern reflects spectral skewness or asymmetry in EEG waveforms, and the d*H* pattern represents stronger inter-regional synchronization during seizure evolution, particularly in focal epilepsy. These findings further support the utility of the d*H* pattern as a non-linear feature in delineating the EZ. Future clinical practices in defining the EZ would benefit from incorporating an integrated EEG biomarker that includes analysis of the d*H* pattern.

We analyzed the morphological features of the *H* pattern in terms of shape and clarity. The shape of the spectral bands in the *H* pattern reflects variations in the frequency of synchronized neural oscillations. The most frequently observed shape is a declining line, which closely resembles a spectral signature known as *chirps* ^33–35^. This shape may indicate a decline in neural firing, possibly due to increased inhibition. In our cohort, nearly half of the SOPs exhibiting low-frequency activity (LVFA) expressed *chirp* ^13^ patterns, with proportions similar to those of the parabolic shape. We propose that some *H* patterns with a parabolic shape and very short upward lines might be classified as chirps due to the limitations of visual inspection. This potential misclassification could narrow the scope of the ictal *H* pattern and limit its applicability. A corticothalamic computational model has similarly elucidated how changes in physiological parameters affect the dynamics of tonic-clonic seizures, similar to the *H* pattern. This model revealed that increasing the connection strength enhances the power, count, and duration of harmonic bands ^36^. Conversely, the clarity of the *H* pattern signifies the distinctness of the harmonic signals against the background. A highly clear *H* pattern is characterized by a narrow frequency band with minimal noise. Interestingly, these morphological properties do not correlate with clinical factors such as seizure-onset pattern, seizure origin, MRI findings, pathological findings, or seizure outcome. The primary clinically relevant aspect of the *H* pattern is its degree of non-linearity. Regardless of its shape, the prominence of the *H* pattern indicates epileptogenicity, and its complete removal has been linked to the potential achievement of seizure freedom.

We observed a stronger association between the d*H* pattern and long-term seizure freedom in neocortical epilepsy compared to mTLE. Previous studies have reported that patients with mTLE generally experience more favorable surgical outcomes than those with neocortical epilepsy ^37^. Additionally, the classification of seizure outcomes can vary among patients. Therefore, we used Engel class Ia as the criterion for a favorable outcome to evaluate the efficacy of the d*H* pattern in localizing the EZ. Earlier iEEG studies have indicated that the extent of SOZ removal does not always correlate with seizure freedom across all patients ^38, 39^. In contrast, we found that, for mTLE, the complete removal of the SOZ had a stronger correlation with long-term seizure freedom than the d*H* pattern. Furthermore, our current mTLE cohort included more MRI-negative cases compared to our previous cohort ^14^. Surgical outcomes in MRI-negative cases are usually less favorable compared to those in TLE-HS ^40, 41^, suggesting a more extensive epileptogenic network in MRI-negative TLE. Previous research has highlighted differences in the involved neural networks between mTLE and neocortical epilepsy. For instance, an ^18^F-FDG-PET study revealed distinct metabolic profiles between these two types of epilepsy, suggesting variations in seizure propagation ^42^. Additionally, an iEEG study showed that the interictal epileptogenic network in mTLE exhibits a more regular pattern of graph connectivity compared to neocortical epilepsy ^43^. It is well-recognized that epileptic networks differ between mTLE and neocortical temporal lobe epilepsy, as well as between TLE and extra-TLE ^44, 45^. A longer follow-up period is necessary to assess the final prognosis in patients with mTLE and to further estimate the value of the *H* pattern in this population.

Nevertheless, our study has several limitations. First, we based the determination of the *H* pattern’s dominance solely on visual interpretation throughout the study. Future investigations should focus on optimizing the definition of the d*H* pattern and developing a more automated algorithm to quantify the *H* pattern. This would enhance the precision and objectivity of the analysis. We plan to create a quantitative index for the *H* pattern to address this issue. Additionally, we did not analyze the occurrence of the second *H* pattern during the later ictal stage of one seizure in this study, which warrants further investigation.

### Conclusions

In focal epilepsy, the dominance of the ictal *H* pattern offers new insights for localizing the EZ, regardless of its morphology. The distinct non-linear feature of the H pattern has significant implications for presurgical assessments and surgical planning. Applying this biomarker in neocortical epilepsy may offer greater value in achieving long-term favorable outcomes compared to mTLE.

## Data availability

The that support the findings of this study are available upon reasonable request from the corresponding author.

## Acknowledgements

This study received support from the National Natural Science Foundation of China (grant number: 82171437, 82301636, 82471469) and the Major Program of the National Natural Science Foundation of Zhejiang Province (grant number: LD24H090003).

## Supplementary material

Supplementary material is available online.

