## Supplemental Table 1-2 and Figure 1-5 for "Dominant harmonic pattern as an ictal marker of the epileptogenic zone in focal neocortical epilepsy"

APPENDIX

Supplementary Material

Number of tables: 2

Number of figures: 5

Table S1. Association between ictal *H* pattern in focal- and non-focal onset epilepsy

|  | Focal-onset  (n = 99) | Non-focal onset  (n = 32) | *p* value |
| --- | --- | --- | --- |
| Ictal *H* pattern, n (%) | 92 (92.9%) | 30 (93.8%) | 1.000 |
| Non-localizable, n (%) | 0 | 6 (20.0%) | **0.0001** |
| Localizable, n (%) | 92 (100%) | 24 (80.0%) |  |
| Two types *H* pattern, n (%) | 0 | 5 (20.8%) | **0.0001** |
| One *H* pattern, n (%) | 92 (100%) | 19 (79.2%) |  |

*H* pattern, *Harmonic* pattern. Notion: If two types of initial ictal *H* pattern were respectively expressed in different seizures originated from each SOZ in one patient, then the patient was considered as having two types of ictal *H* pattern.

Table S2. Association between morphology of ictal *H* pattern and clinical characteristics

|  | Grade | | *p* value | Shape | | | *p* value | Type | | | | | | *p* value |
| --- | --- | --- | --- | --- | --- | --- | --- | --- | --- | --- | --- | --- | --- | --- |
|  | A | B |  | 1 | 2 | 3 |  | A1 | A2 | A3 | B1 | B2 | B3 |  |
| Number of patients, n | 57 | 60 | - | 47 | 67 | 3 | - | 22 | 34 | 1 | 25 | 33 | 2 | - |
| Lesional MRI, n (%) | 36 (63.2%) | 41 (68.3%) | 0.555^a^ | 33 (70.2%) | 42 (62.7%) | 2 (66.7%) | 0.704^b^ | 15 (68.2%) | 20 (58.8%) | 1 (100%) | 18 (72.0%) | 22 (66.7%) | 1 (50.0%) | 0.811^b^ |
| Focal-onset, n (%) | 45 (78.9%) | 47 (78.3%) | 0.935^a^ | 40 (85.1%) | 50 (74.6%) | 2 (66.7%) | 0.347^b^ | 17 (77.3%) | 27 (79.4%) | 1 (100%) | 23 (92.0%) | 23 (69.7%) | 1 (50.0%) | 0.261^b^ |
| Neocortex-onset, n (%) | 28/45 (48.9%) | 33/47 (51.1%) | 0.418^a^ | 28/40 (70.0%) | 31/50 (62.0%) | 2/2 (100%) | 0.317^b^ | 11/17 (64.7%) | 16/29 (59.3%) | 1/1 (100%) | 17/23 (73.9%) | 15/23 (65.2%) | 1/1 (100%) | 0.719^b^ |
| Pathology, n (%) | 46 | 53 | 0.565^b^ | 41 | 56 | 2 | 0.195^b^ | 17 | 28 | 1 | 24 | 28 | 1 | 0.397^b^ |
| HS | 8 (17.4%) | 7 (13.2%) |  | 5 (12.2%) | 10 (17.9%) | 0 |  | 3 (17.6%) | 5 (17.9%) | 0 | 2 (8.3%) | 5 (17.9%) | 0 |  |
| FCD type-II | 16 (34.8%) | 14 (26.4%) |  | 15 (36.6%) | 14 (25.0%) | 1 (50.0%) |  | 6 (35.3%) | 9 (32.1%) | 1 (100%) | 9 (37.5%) | 5 (17.9%) | 0 |  |
| Other MCD types | 7 (15.2%) | 14 (26.4%) |  | 10 (24.4%) | 11 (19.6%) | 0 |  | 4 (23.5%) | 3 (10.7%) | 0 | 6 (25.0%) | 8 (28.6%) | 0 |  |
| Tumor | 2 (4.3%) | 1 (1.9%) |  | 0 | 2 (3.6%) | 1 (50.0%) |  | 0 | 2 (7.1%) | 0 | 0 | 0 | 1 (100%) |  |
| Gliosis/other non-specific | 13 (28.3%) | 17 (32.1%) |  | 11 (26.8%) | 19 (33.9%) | 0 |  | 4 (23.5%) | 9 (32.1%) | 0 | 7 (29.2%) | 10 (35.7%) | 0 |  |
| Engel Ia, n (%) | 25/49 (51.0%) | 32/54 (59.3%) | 0.401^a^ | 24/42 (57.1%) | 31/59 (52.5%) | 2/2 (100%) | 0.271^b^ | 10/18 (55.6%) | 14/30 (46.7%) | 1/1 (100%) | 14/24 (58.3%) | 17/29 (58.6%) | 1/1 (100%) | 0.625^b^ |

a. *Chi-Squared* test. b. *Fisher exact* test.

H pattern, Harmonic pattern, HS, hippocampal sclerosis, MRI, magnetic resonance imaging, FCD, focal cortical dysplasia, MCD, malformation of cortical development

Notion: Only patients expressing one ictal *H* pattern were enrolled in the analysis. Pathological results of six patients were lacking because they had received laser ablation as

treatment.

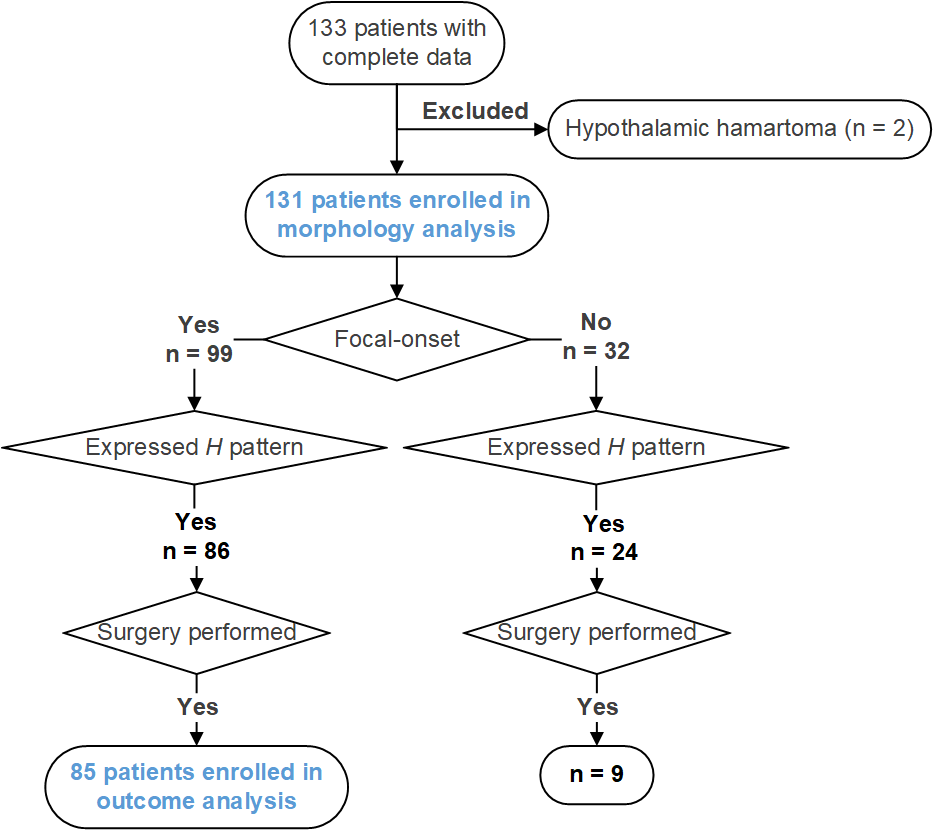

**Figure S1.** A flowchart demonstrating the inclusion and exclusion criteria of the patients enrolled in the morphology analysis and outcome analysis.

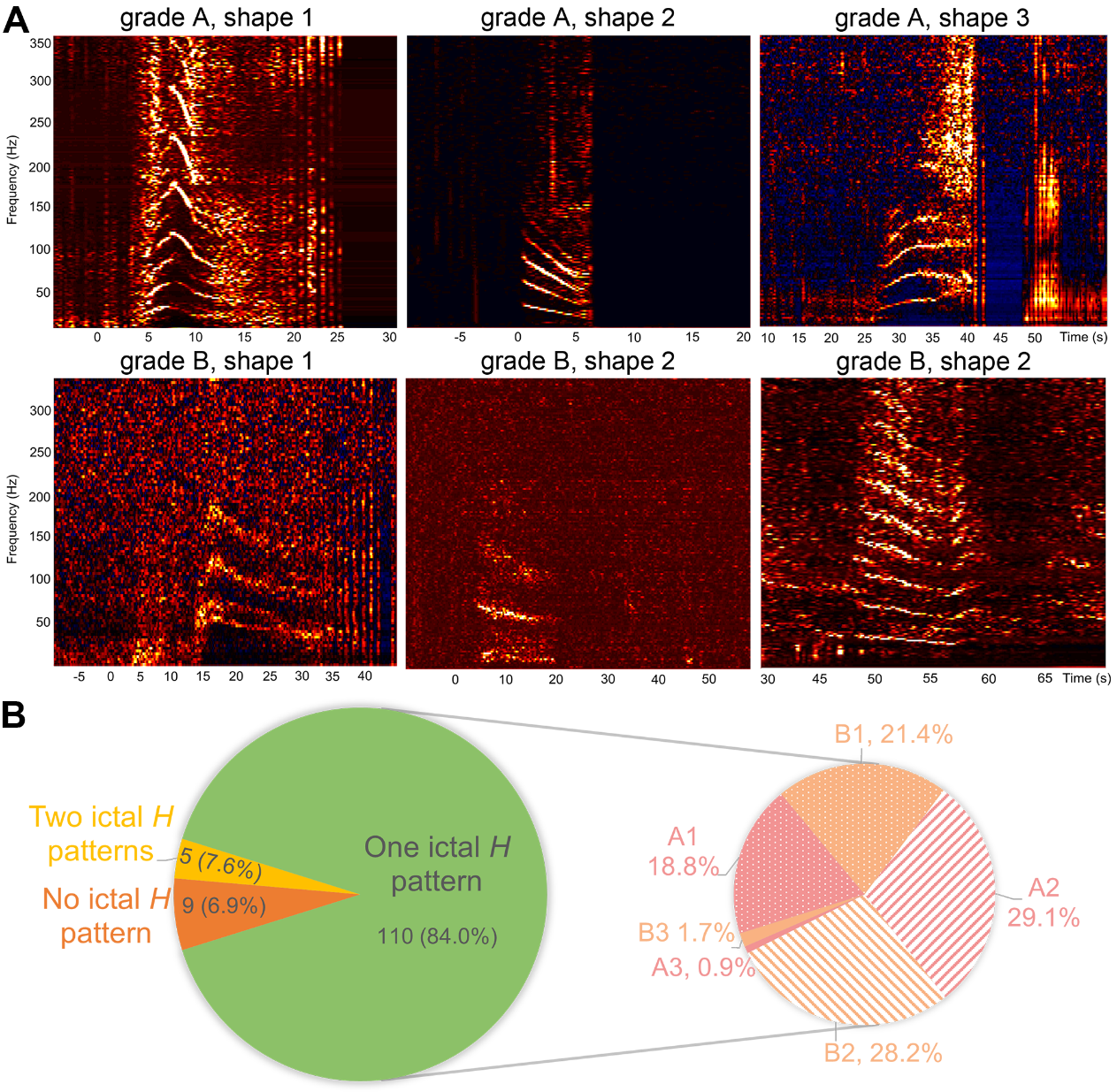

**Figure S2.** **Schematic diagrams and distribution of different morphology of ictal *H* pattern.** Five morphological types are shown (A). In the upper row, three time-frequency (TF) plots exhibiting a clear clarity border represent three different shapes of the spectral bands, which are ‘parabola’, ‘declining line’, ‘ascending line’ (from the left to right). In the bottom row, three time-frequency (TF) plots exhibiting a fuzzier border represent two different shapes of the spectral bands, which are ‘parabola’, ‘declining line’, ‘declining line’ (from the left to right). (B) The left pie chart showed the distribution of expressing one ictal *H* pattern (green) or two ictal *H* patterns (yellow), and no ictal *H* pattern (orange) in the morphology cohort. The right pie chart exhibited the distribution of six morphological types in patients expressing one ictal *H* pattern. Grade A and B are represented with pink color and light orange color. Shape 1 and 2 are illustrated by dots and lines wrapped by a scallop shape respectively. Shape 3 is displayed by a full-colored scallop shape.

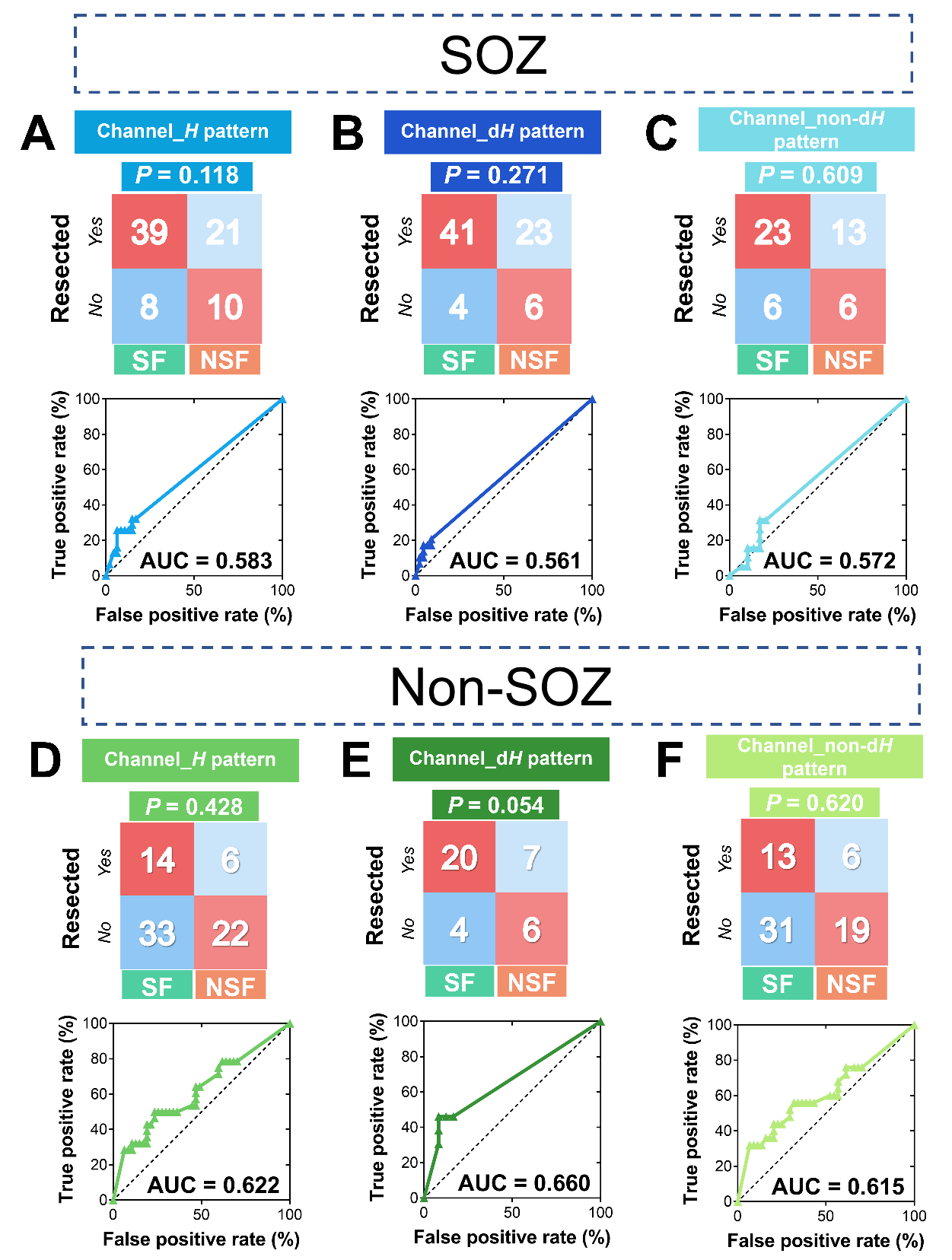

**Figure S3.** **Predictive values for seizure outcome.** Four-fold tables and receiver operator characteristic curve (ROC) for resected ratios of channels expressing ictal *H* pattern (bright blue; bright green), channels expressing ictal dominant *H* patten (dark blue; dark green) and channels expressing ictal non-dominant *H* pattern (light blue; light green) within SOZ (A-C) and within non-SOZ (D-F) as predictor of the post-surgical outcome. In the four-fold tables above, the numbers of seizure-free patients and non-seizure free patients who had the corresponding channels all removed (represented by ‘*Yes’* listed on the upper left of the table), or incompletely/none removed (represented by ‘*No*’ listed on the bottom left of the table) were respectively listed in the box. Of each *Chi-Squared* test, the *p* value was listed above the table. In the ROC curves (continuous colored line), with the black dashed lines representing the diagonal of each curve, each colored dot represents a value combined by the false positive rate on X-axis and the true positive rate on Y-axis. The areas under curves were respectively listed on the bottom right corner of the curves.

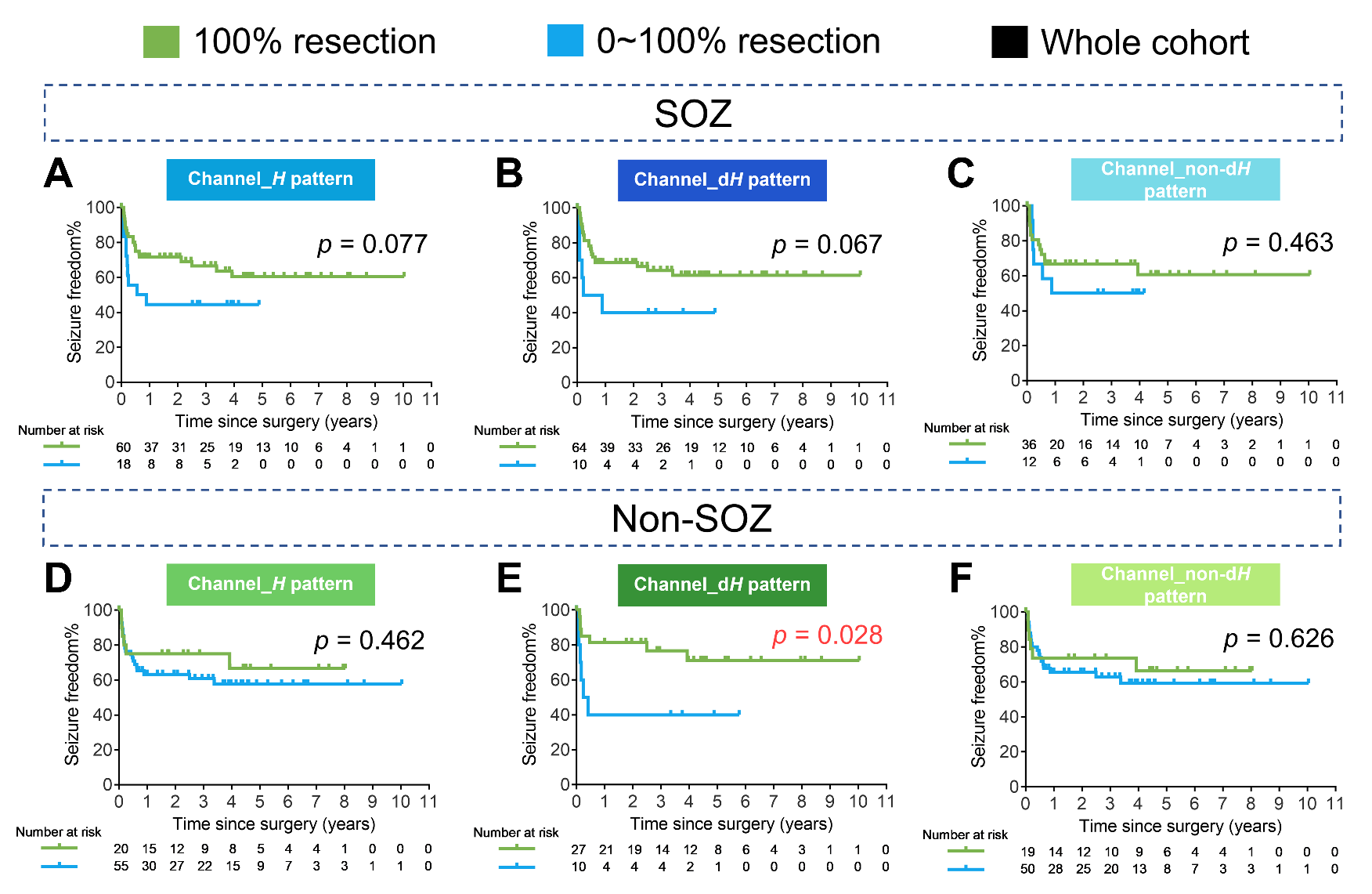

**Figure S4.** **Survival analysis.** Kaplan-Meier (KM) plots of cumulative probability of continuous seizure freedom separated by complete (100%) resection (green continuous line) and incomplete/no (0~100%) resection (blue continuous line) of channels expressing ictal *H* pattern, channels expressing ictal dominant *H* patten and channels expressing ictal non-dominant *H* pattern within SOZ (A-C) and within non-SOZ (D-F). The ‘number at risk (number censored)’ at each year is annotated below the KM plot for every category.

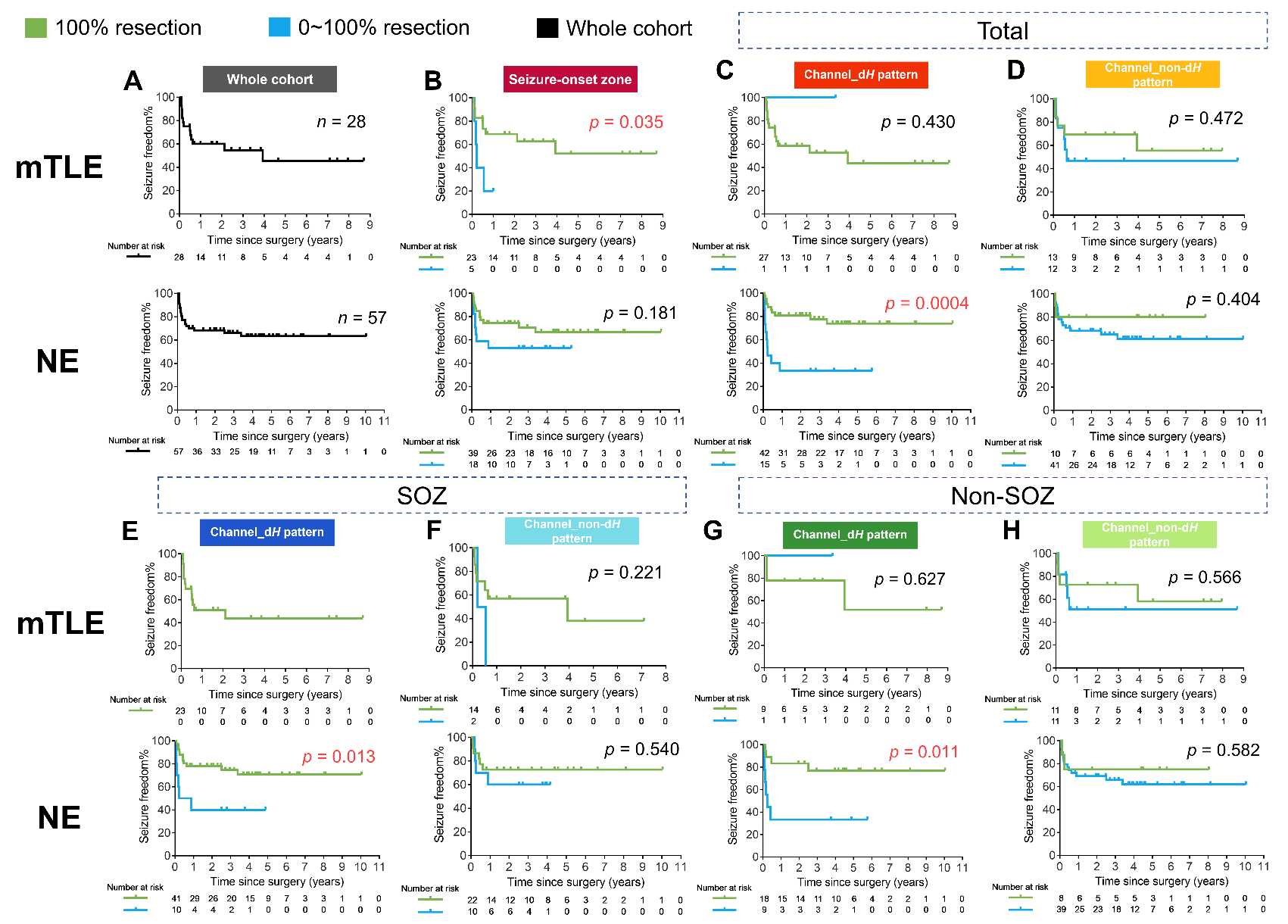

**Figure S5.** **Survival analysis in relation to epilepsy type.** Kaplan-Meier (KM) plots of cumulative probability of continuous seizure freedom in the (A) mTLE group and (B) neocortical epilepsy group. Kaplan-Meier (KM) plots of cumulative probability of continuous seizure freedom separated by complete (100%) resection (green continuous line) and incomplete/no (0~100%) resection (blue continuous line) of (B, F) SOZ, (C, G) channels expressing ictal dominant *H* patten and (D, H) channels expressing ictal non-dominant *H* pattern. The results were further classified into the mTLE group and neocortical epilepsy group, respectively. The ‘number at risk (number censored)’ at each year is annotated below the KM plot for every category.
